# Motor and Activity Psychosis-Risk (MAP-R) Scale: An exploration of scale structure with replication and validation

**DOI:** 10.1101/2020.07.02.20145482

**Authors:** Katherine S. F. Damme, Jason Schiffman, Lauren M. Ellman, Vijay A. Mittal

**Author notes:** **Corresponding Author:** Katherine Damme, Department of Psychology, Northwestern University, 2029 Sheridan Rd., Evanston, IL 60208.

## Abstract

**Background:** Motor abnormalities precede and predict the onset of psychosis. Despite the practical utility of motor abnormalities for early identification, prediction, and individualized medicine applications, there is currently no dedicated self-report instrument designed to capture these important behaviors. The current study assessed and validated a questionnaire designed for use in individuals at clinical high-risk for psychosis (CHR).

**Methods:** The current study included both exploratory (n=3,009) and validation(n=439) analytic datasets– that included individuals at CHR (n=84)-who completed the novel Motor Abnormalities and Psychosis-Risk (MAP-R) Scale, clinical interviews and a finger tapping task. The structure of the scale and reliability of items were consistent across two analytic datasets. The resulting scales were assessed for discriminant validity across CHR, community sample non-psychiatric volunteer, and clinical groups.

**Results:** The scale showed a consistent structure across independent data points across two analytic datasets subscale structure. The resultant subscale structure was consistent with conceptual models of motor pathology in psychosis (coordination and dyskinesia) in both the exploratory and the validation analytic dataset. Further, these subscales showed discriminant, predictive and convergent validity. The motor abnormality scales discriminated CHR from community sample non-psychiatric controls and clinical samples. Finally, these subscales predicted to risk calculator scores and showed convergent validity with motor performance on a finger tapping task.

**Conclusion:** The MAP-R scale demonstrated good internal validity, discriminant validity, predictive validity, and convergent validity, and subscales map on to conceptually relevant motor circuits. This scale showed great promise in characterizing a novel area of detection of psychosis risk.

## Introduction

Motor abnormalities have been found to precede and predict the onset of psychosis.^1–8^ These motor abnormalities are tied to cognitive deficits, functional outcomes and other core features of psychotic disorders.^9–14^ Taken together with a growing body of supporting structural,^15–18^ connective, and functional imaging results,^19–21^ imply that motor features reflect important vulnerability and disease driving mechanism.^11–13,22^ Relatedly, motor symptoms may be useful in forming distinct clinical high-risk (CHR) subtypes.^19^ Motor behaviors may also useful in monitoring side effects,^23^ promoting treatment planning,^24^ and tracking treatment outcome.^25^ In addition to motor abnormalities, sedentary behavior and level of aerobic activity are also important risk/protective indicators for individuals at CHR. ^26–29^ Despite the clear importance of motor symptoms and activity, at the present time there is no dedicated self-report measure for assessing them in psychosis risk populations. Current motor abnormality measures often require clinical training and equipment,^30,31^ making current testing may be impractical for widespread clinical use. Additionally, extant self-report and clinical motor assessments tend to assess a limited number of motor abnormalities, but psychotic disorders are characterized by numerous motor abnormalities^31^ of clinical importance (e.g. conversion, subtyping).^1,19,32^ This paucity of motor abnormality items in clinical assessments limits the power to detect relevant motor abnormalities related to CHR symptoms, conversion, and subtyping.

Motor abnormalities have been found to reflected an early vulnerability to individuals at CHR for psychosis. Studies of infants that later develop psychosis in adulthood suggest that coordination deficits, delays, and dyskinesias characterize an early vulnerability.^3^ Similar work in people at CHR suggested that coordination deficits and delays in motor learning reflect cerebellar circuit dysfunction,^17,21,33^ whereas slowing as well as hyperkinetic movements reflect basal ganglia circuit pathology.^15^ Notably, these circuits are relevant to prominent conceptual theories of psychosis,^22,34^ and relatedly, both domains predicted worsening course,^1,2,21^ poor functional outcomes,^35,36^ and ultimate conversion to psychosis.^1,32,37–39^ Further, neuroimaging studies in individuals at CHR have refined our understanding of neural underpinnings, as well as the links with disease-relevant mechanisms.^40–45,9,15,46–49 22,43,50,51^ With respect to physical activity, individuals at CHR show decreased levels of physical activity;^26,28,29,52^ level of physical activity has been linked to abnormalities in the hippocampus among other regions.^26,53–55^ Indeed, exercise interventions aimed at increasing physical activity have shown evidence of increased cognition, reduced symptoms, and improved functional connectivity in the hippocampus.^27^ Due to the predictive quality, motor abnormalities and physical activity may be particularly useful to assess psychosis risk.

Despite its promise motor functioning has been largely under-utilized in the early assessment of psychosis risk; some assessments include no motor items^56,57^ and others only including one^58,59^ or two items.^59,60^ Indeed motor abnormalities are suggested to be a unique and independent feature of psychosis risk.^11–13,22,42,61^ Thus, adding a motor measure to the standard psychosis risk assessment may increase sensitivity and provide added benefit to prediction algorithms for conversion to psychosis. The current study developed a novel Motor Abnormalities and Psychosis-Risk (MAP-R) Scale. First, we examined structure of responses replicated across two analytic datasets; the exploratory dataset -a larger (n=3,009) screening analytic dataset who completed the MAP-R scale (as part of a larger non-motor battery) and only completed phase one of the study- and the scale validation analytic dataset-a smaller independent data points that completed additional validation measures during phase two of the study (n=439). Then we reported the psychometrics statistics (reliability) of the resulting subscales established by the response structure across both analytic datasets. The resultant subscales were used in subsequent analyses to probe the discriminant, predictive, and convergent validity of these scales. Discriminant validity was examined by assessing the degree to which these scales differed between relevant CHR groups compared to other disorders (depression and anxiety) and a community control sample. Convergent validity was assessed by relating scale items to motor performance during a motor neurocognitive task (i.e., finger tapping task). Finally, to examine predictive validity, these scales were related to the calculated risk for psychosis (SIPS-RC).^20,62^

## Methods and Materials

### Participants

Participants were recruited as a part of a large, multi-site community sample known as the Multisite Assessment of Psychosis-Risk study. The primary focus of that study was to evaluate markers of risk for psychosis in a large, representative community sample across multiple study sites. Study sites included the greater catchment areas of Philadelphia (Temple University), Chicago (Northwestern University), and Baltimore (University of Maryland-Baltimore County). Recruitment occurred through various outlets including ads on various internet sites (e.g., Craigslist, Facebook), student volunteer pools, refer-a-friend links, and flyers. Recruitment centered on non-clinical sources, therefore no recruitment took place at clinical locations such as outpatient psychiatric clinics and hospitals in an attempt to keep the community sample relatively unbiased.

### MAP Study – Screening Phase

The study contained two phases, *Figure 1*. The first phase was completed by all 3,448 participants and included an online battery of measures, including established psychosis risk questionnaires-e.g., the prodromal questionnaire^56^ (PQ), the PRIME Screen^58^-and a variety of other questionnaires, including the Motor Abnormalities Psychosis Risk Scale. The first phase identified individuals at probable high or probable low-risk for psychosis. Notably the sites did not differ in sample demographics, such as biological sex at birth (*p*=.19) or age (*p*=.38). A random sample balanced across probable low and probable high-risk groups was invited for the second phase. Participants were labeled as questionnaire high-risk for psychosis if they endorsed eight or more distressing items on the PQ and two items with a distress rating of 5 or one item as a 6 on the PRIME scale. Subjects who did not meet either threshold were randomly selected from below these cut-offs and treated as questionnaire low-risk.The second phase also had additional inclusion criteria including participants must be proficient in English, between the ages of 16 to 30, and have normal or corrected to normal vision. Additional exclusion criteria included either an inability to provide consent for the second phase or an unwillingness to attend and in-person study visit. For the current analyses, the presence of a current psychotic diagnosis (n=3, as assessed during an in-person visit) was also an exclusion criterion. To preserve the representativeness of the community sample, there were no other exclusion criteria.

**Figure 1.**
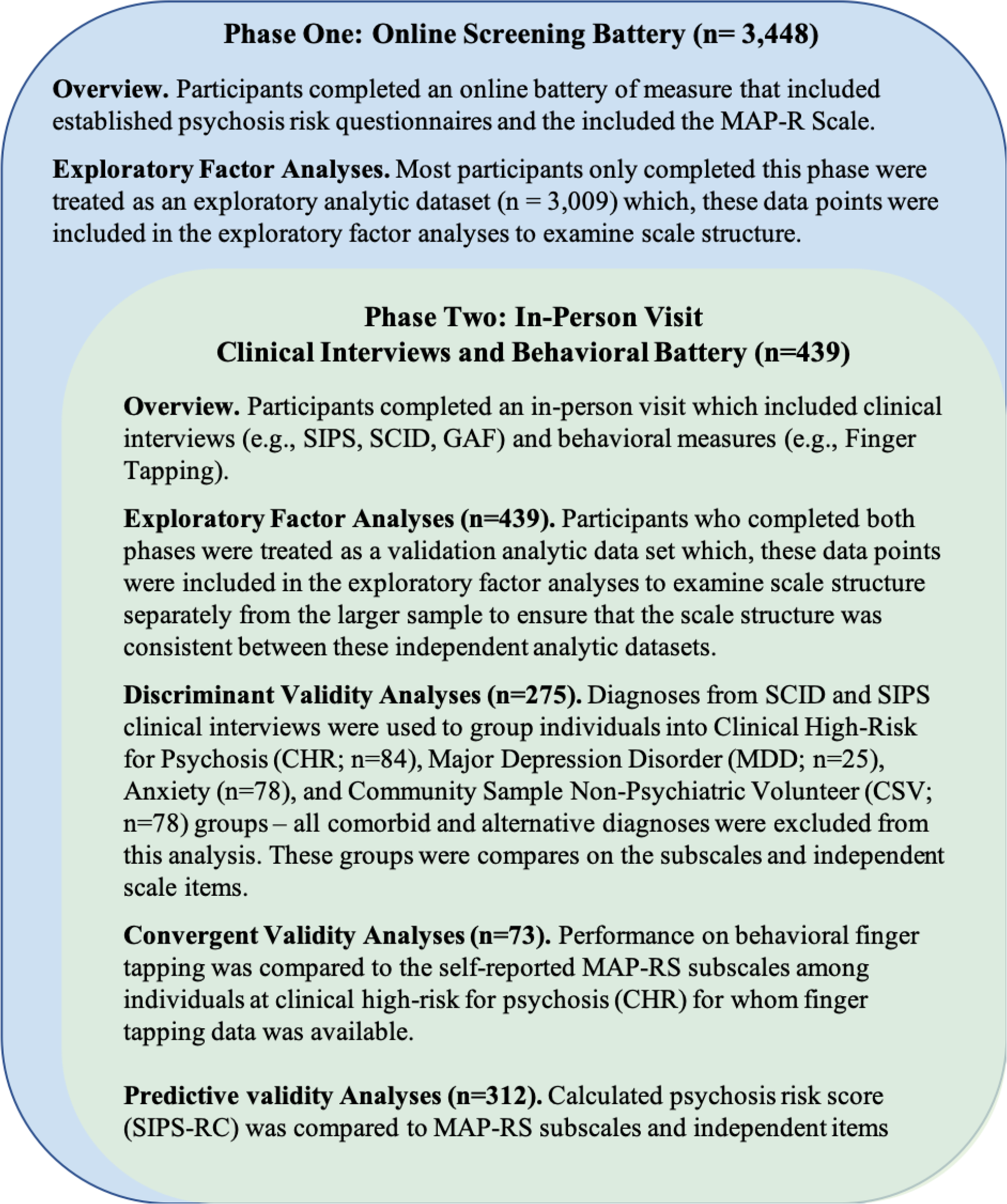
Study Sample: Guide on data collection phases, sample recruitment overlap, and measure sample sizes.

### MAP Study – Validation Phase

The second, validation phase was an in-person visit at the university study site that included a variety of relevant and discriminant measures to validate the current scale. These validation measures were collected within a larger study battery that included clinical assessments using both the Structured Interview for Psychotic Risk^58^ (SIPS) and the Structured Clinical Interview for DSM-5 Research Version^60^ (SCID-5-RV). During the clinical assessment individuals were classified as clinical high-risk for psychosis (CHR) if they met one of the following criteria: attenuated positive symptoms or genetic risk and deterioration of function. Attenuated positive symptom criteria was determined by the SIPS guidelines. Finally, genetic risk and deterioration of function required that a participant have a first degree relative with a diagnosis with a psychotic disorder or schizotypal personality disorder and that reported that they experienced functional decline in the past year, resulting in 85 individuals being classified as CHR. In addition to assessing the presence of psychosis or CHR status, the SCID provided comorbid diagnoses across the entire community sample. The alternate diagnoses in the community sample was used to assess whether MAP-R subscales and items can discriminate between CHR status and other psychopathology detected in the community validation analytic dataset. The second phase included the completion of the Penn computerized neurocognitive battery and additional questionnaires. There were 13 individuals with lifetime antipsychotics use, which included individuals from the anxiety (n=4; 5.2%), CHR (n=8, 9.5%), and depression group (n=1, 2.9%).

### Motor Abnormalities Psychosis Risk Scale

The Motor Abnormalities Psychosis Risk (MAP-R) Scale is a 14-item questionnaire that included questions about early developmental motor delays, the frequency of abnormal motor experiences, general assessments of motor function, and frequency of physical activity. Questions were developed based on a review of extant literature^12,22,31,41,61^ regarding motor abnormalities in individuals at CHR to include features of motor abnormalities linked to symptoms,^4,10,19,21,48,48,63^ conversion,^1,5,32,37,38^ and clinical/cognitive subtyping^19^ of psychosis risk. From this review of the literature a set of items were compiled to examine relevant constructs (e.g., ballistic movements, sway/balance). These items included several likert-type scale responses, such as “On a scale of 1-10, how would you rate your current motor skills?” and “Do you enjoy hobbies where you use skilled hand movements (e.g., art, crafts)? (Responses: Never Sometimes Lots)”. For an idealized version of the final measure and scoring, see Supplemental Materials.

### SIPS Risk-Calculator

The SIPS-RC is a Structured Interview on Prodromal Risk Symptoms (SIPS) -based risk calculator that provided practical, individualized risk assessments that can be easily implemented in a clinical setting.^62,64^ In this calculator, risk probability assessments are derived from four dimensions: positive and negative symptom severity, deterioration of function over the past 12-months, and low levels of dysphoric mood. In the current study, the global functioning scale^65^ was used to assess functional decline in the past year. Positive and negative symptom severity was assessed during the SIPS interview, and quantified as a composite score of endorsed positive and negative symptoms.^64^ Finally, low levels of dysphoric mood were assessed during the general symptom assessment of the SIPS.^62,64^ This approach has been validated against external independent risk calculators.^62,64^

### Finger Tapping - Motor Validation Task

In the current study we examined the performance on the computerized finger tapping (CTAP) performance.^66^ During the task, participants were instructed to press the spacebar as quickly as possible moving only the index finger for 10 seconds after their first button press. In this task, trial blocks alternated between the dominant and non-dominant hand for 10 blocks (5 block per hand). Median finger tapping and tapping variability behavior was used as a convergent measure of motor performance to assess the sensitivity of the MAP-R scale to motor performance.

### Analytical Strategy

Exploratory factory analyses were used to examine structure of responses in the two analytic datasets (i.e., exploratory and validation). These two analytic datasets, with independent data-points include a larger exploratory, screening analytic dataset (n=3,109) that completed only phase one of the study and the scale validation analytic dataset (n=439) that completed both phase one and phase two.^67^ For each analysis, all participants with the data for that given analyses were included, rather than reducing the study sample size to only those individuals with a full and complete dataset; the intention in this approach was to both maximize power and transparency, *see Figure 1 for analyses sample sizes*. To assess the appropriate number of factors for these analyses, two approaches were used: 1) the Cattell’s scree test was used to identify the appropriate number of factors in the correlation matrix, 2) the eigen values were graphed against the increasing number of factors and components with both a threshold provided by simulated data. Factor analyses in both the exploratory analytic dataset (only completing the MAP-R scale among a battery of non-motor initial screening scales) and the validation analytic dataset (who completed both study phases) were used to determine the optimal items to composite into subscales. Then, we reported the psychometrics statistics, including internal consistency statistics (Cronbach’s alpha and Guttman’s Lambda) for the resulting subscales. Factor analyses and item reliability was assessed using the R v.4.0.0^68^ and the Psych package.^69^

All items that were grouped into a factor were examined as a composite subscale in the subsequent analyses. Items that were not included in the subscales (i.e. items that are not related to other items) were treated as independent items and evaluated for discriminant validity and convergent validity. Independent items that failed to show discriminant and convergent validity were excluded from the final scale. These subsequent analyses probed the discriminant and predictive validity of these composite subscales and reduced the total number of comparisons.

To examine discriminant validity, we examined the degree to which these scale factors differ between relevant CHR group (n=84) compared other disorders, i.e., depression (n=35) and anxiety (n=78), and a CSV (n=78) samples among the validation analytic dataset, using general linear models for the composite scales and chi-square for independent items. To examine predictive validity, these scale factors were related to the calculated risk for psychosis (SIPS-RC) and current symptom severity (SIPS total scales) within the CHR group from the validation analytic dataset. To examine convergent validity, the motor scales were compared to the motor performance (i.e., finger tapping task) in the CHR validation analytic dataset. All validation analyses were completed in SPSS version 26.

## Results

### Participants

There were no significant differences in key demographic variables including biological sex at birth, ethnicity, race, age, and income were compared across exploratory and validation analytic datasets, as well as discriminatory clinical samples, Table 1.

**Table 1.**
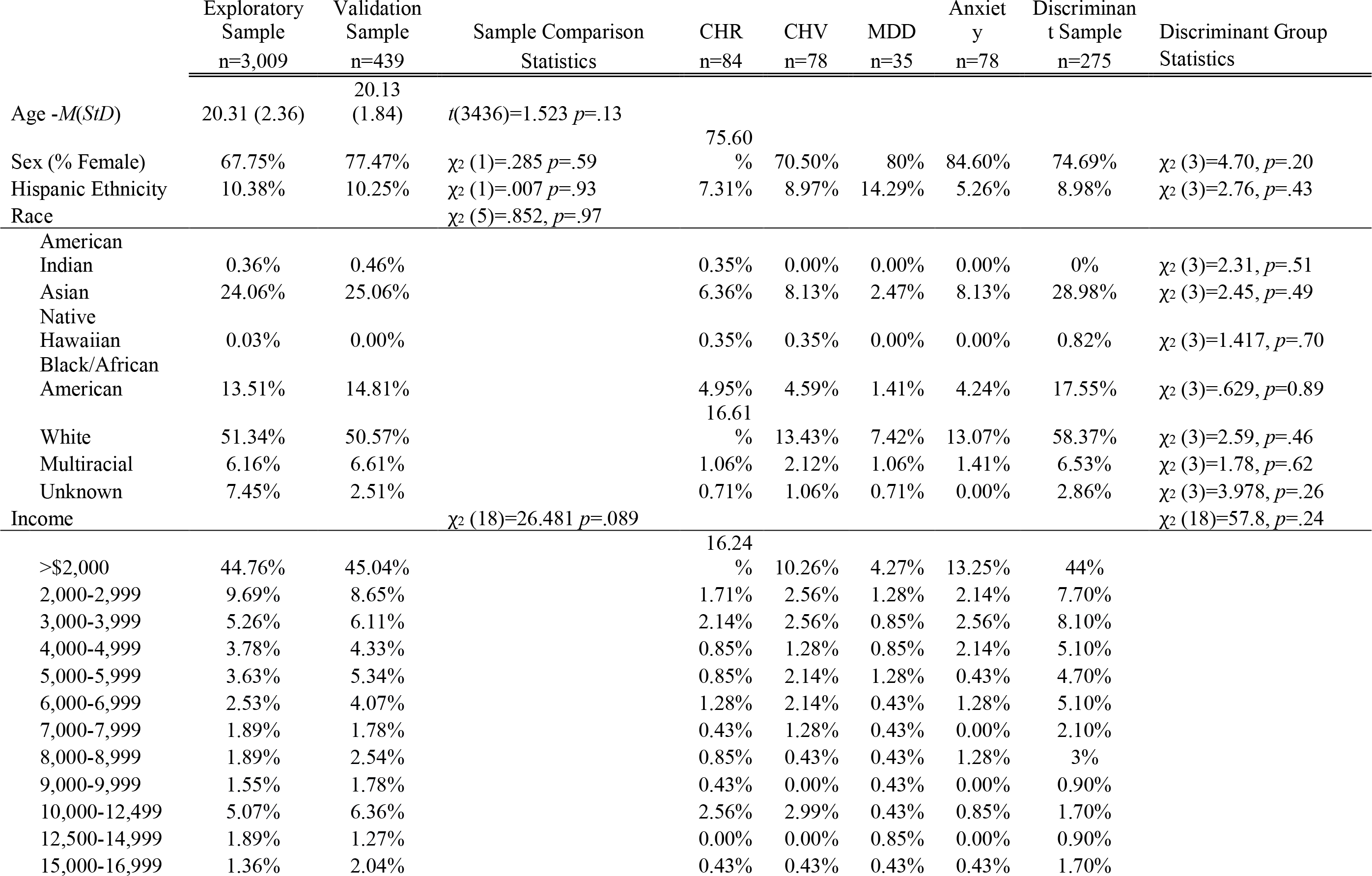
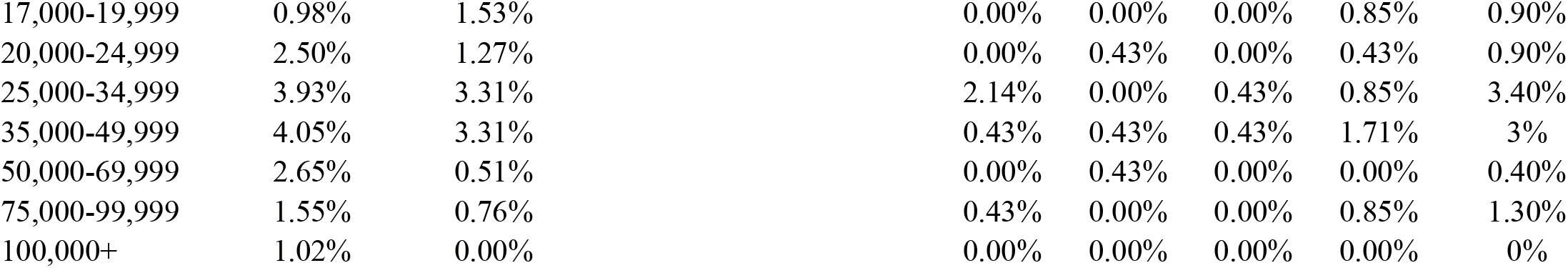
Demographic Metrics: A comparison of diagnostic groups within the validation sample for discriminatory analyses

### Exploratory factor analyses

In the exploratory analytic dataset, the traditional Cattell’s scree analyses suggested that the intercorrelation matrix showed 4 components; scree analyses compared to simulated data suggested the presence of 3 components or 2 factors, *Supplemental Figure 1*. Both analytic datasets showed similar scale structures with 2 components dividing the scale into ***Motor Abnormality*** items and ***Physical Activity*** items; when reorganized into three factors, the ***Motor Abnormality*** items divided into sub-facets *coordination abnormalities* items and *dyskinesia* items. In the validation analyses, the traditional Cattell’s scree analyses suggested that the intercorrelation matrix showed 3 components; scree analyses compared to simulated data suggested the presence of 3 components or 2 factors, *Figure 2*. There were two items that did not load onto these factors including an item on motor delays (i.e., “Are you aware of any delays in walking, talking, or toilet training when you were an infant or toddler?”) and an item on fine motor hobbies (i.e., “Do you enjoy hobbies where you use skilled hand movements (art, crafts)?”). These items were treated as independent items in the following validation analyses. Both exploratory and validation analytic datasets showed similar internal consistency (*Table 2*).

**Table 2.** Reliability of scales replicated across samples

| Factor Scales | Exploratory Sample |  | Validation Sample |  |
| --- | --- | --- | --- | --- |
|  | Cronbach's Alpha | Guttman's Lambda | Cronbach's Alpha | Guttman's Lambda |
| Motor Abnormalities | 0.69 | 0.71 | 0.71 | 0.73 |
| Coordination Scale | 0.71 | 0.68 | 0.75 | 0.71 |
| Dyskinesia Scale | 0.67 | 0.51 | 0.68 | 0.51 |
| Activity Scale | 0.68 | 0.64 | 0.69 | 0.64 |

**Figure 2.**
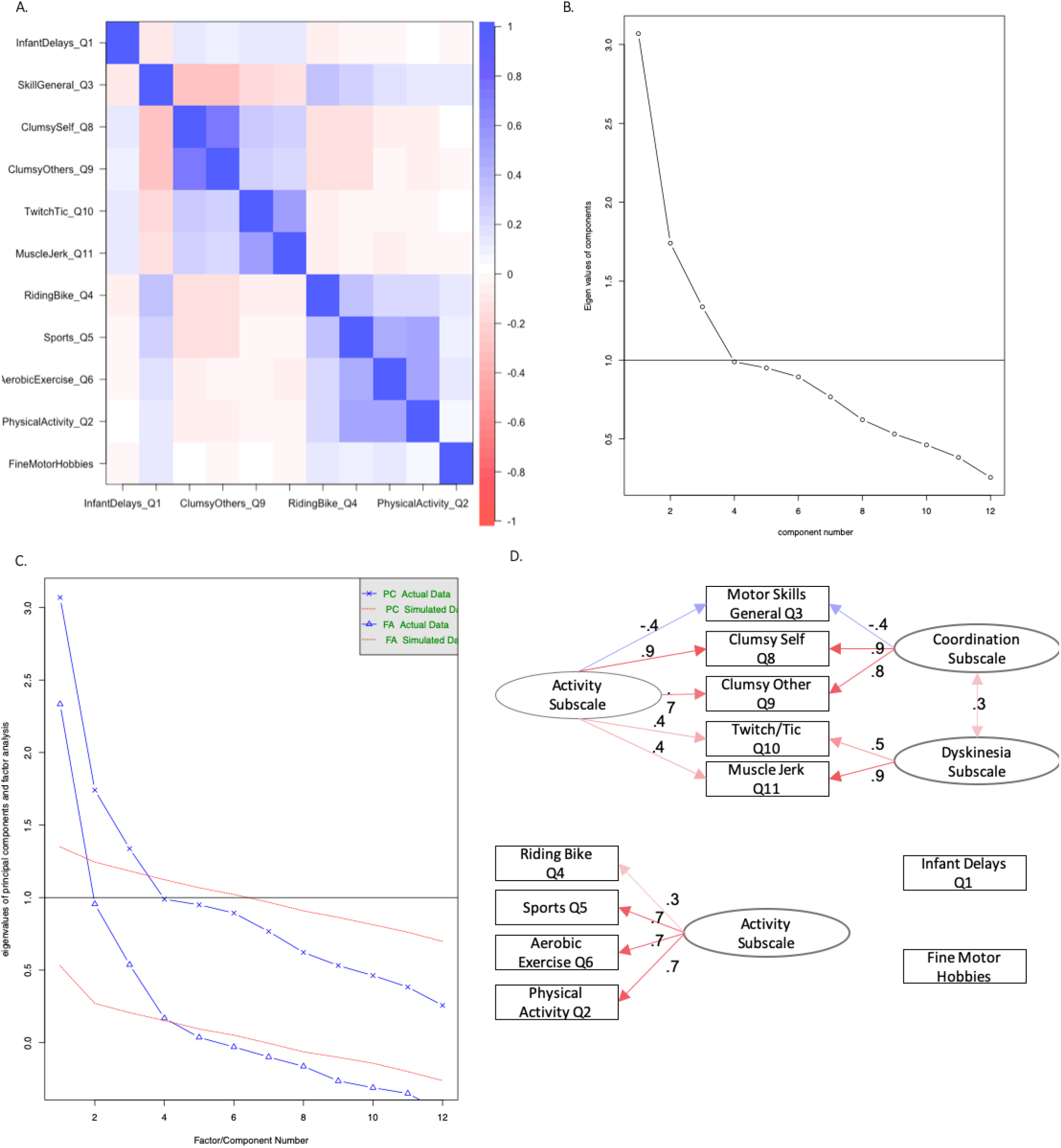
Factor analyses and data structure for the validation analytic dataset: A. Depicts the intercorrelation structure of the items, B. Depicts the Cattell scree plot of the intercorrelation matrix, C. Depicts the factor analyses compared to simulated data in a scree plot, D. Depicts the factor structure for both a 2 factor (***Motor Abnormalities*** and ***Physical Activity Scales***) and three factor solution (*Dyskinesia Abnormalities sub-facets, Coordination abnormalities sub-facets*, and ***Physical Activity Scale***)

### Discriminant validity among clinical samples

Composite subscales totals were compared across diagnostic groups in independent analyses of variance model (ANOVA), *Figure 3*. For each of the independent items a limited set of Likert-responses were compared across groups using a chi-square analysis.

**Figure 3.**
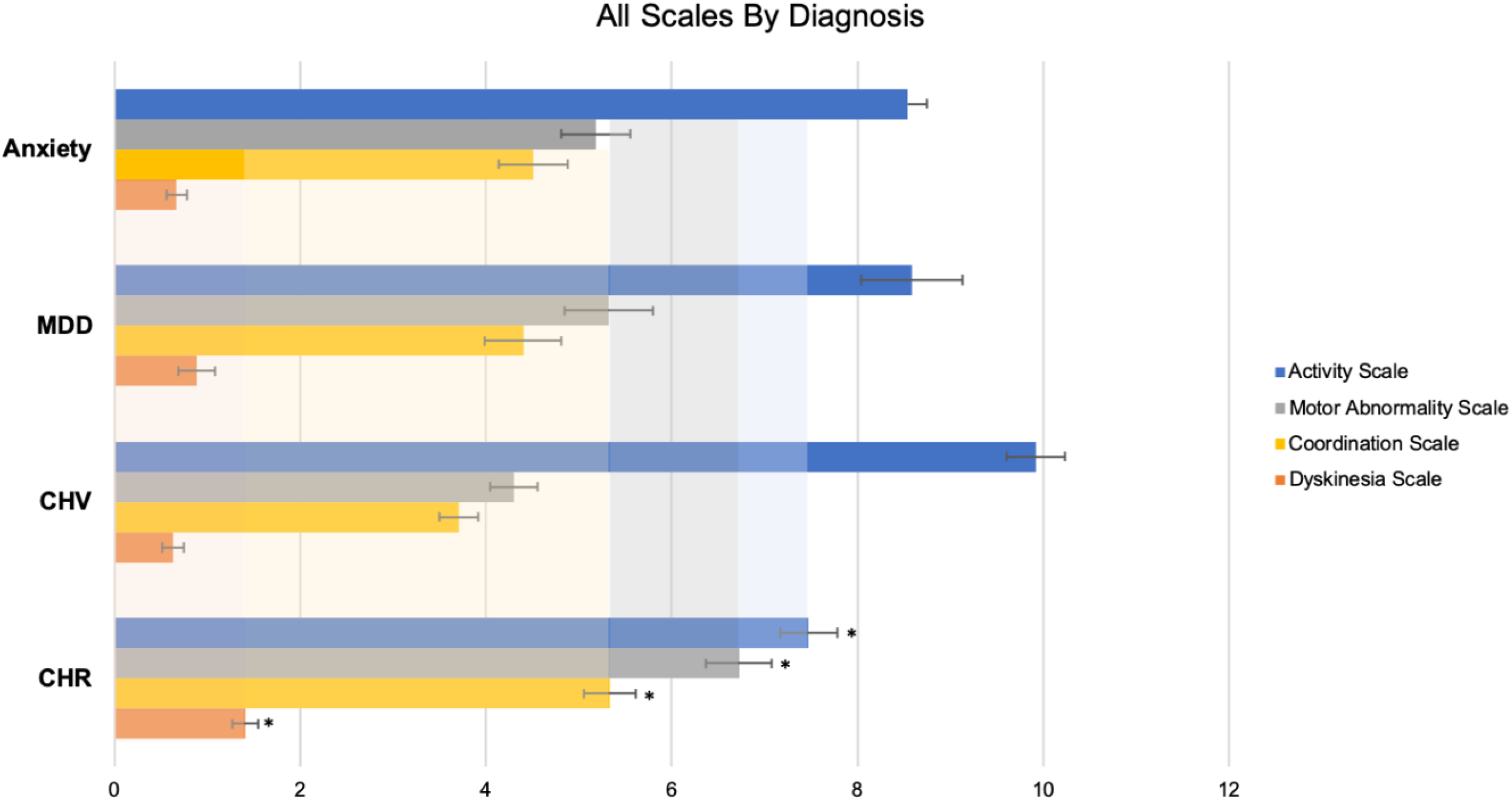
Discriminant Validation Analyses: Composite Scales by Group

#### Motor Abnormalities Subscale by Diagnostic Group

The ***Motor Abnormality*** subscale significantly differed among groups, *F*(270,3)=9.50, *p*<.001, *partial* η^2^*=*.10, such that CHR (*M*=6.72, *StD=*3.25, *SEM*=.36) showed increased ***Motor Abnormality*** score than individuals with depression (*M*=5.32, *StD=*2.78, *SEM*=.37, *p*=.02), anxiety (*M*=5.18, *StD=*3.20, *SEM*=.37, *p*=.006), and the CSV (*M*=4.29, *StD=*2.21, *SEM*=.25, *p*<.001),

#### Abnormalities Sub-facets by Diagnostic Group

The *Coordination Abnormalities* sub-facet significantly differed among groups, *F*(271,3)=6.193, *p*<.001, *partial* η^2^*=*.07, such that CHR (*M*=5.33, *StD=*2.55, *SEM*=.28) showed increased *Coordination Abnormalities* scores than individuals with depression (*M*=4.40, *StD=*2.44, *SEM*=.412, *p*=.05), anxiety (*M*=4.51, *StD=*2.74, *SEM*=.317, *p*=.043), and the CSV (*M*=3.70, *StD=*1.83, *SEM*=.21, *p*<.001). The *Dyskinesia* sub-facet significantly differed among groups, *F*(270,3)=8.54, *p*<.001, *partial* η^2^*=*.09, such that CHR (*M*=1.41, *StD=*1.26, *SEM*=0.14) showed an increased *Dyskinesia* score than individuals with depression (*M*=0.88, *StD=*1.15, *SEM*=0.20, *p*=.02), anxiety (*M*=0.67, *StD=*0.96, *SEM*=0.11, *p*<.001), and the CSV (*M*=0.636, *StD=*1.01, *SEM*=0.12, *p*<.001).

#### Physical Activity Subscale by Diagnostic Group

The ***Physical Activity*** subscale significantly differed among groups, *F*(250,3)=8.536, *p*<.001, *partial* η^2^*=*.09, such that CHR (*M*=7.47, *StD=*2.67, *SEM*=0.31) showed less physical activity than the CSHV (*M*=9.19, *StD=*2.74, *SEM*=0.32, *p*<.001). Although the CHR group showed less physical activity than individuals with depression (*M*=8.59, *StD=*3.18, *SEM*=0.48, *p*=.076) and anxiety (*M*=8.19, *StD=*3.53, *SEM*=0.45, *p*=.16); this difference was not significant.

#### Independent Items by Diagnostic Group

The motor delay response (no delays, some delays, a lot of delays) were compared across the diagnostic groups (CHR, CSV, depression, anxiety); groups differed in terms of motor delays χ^2^(223)=9.45, *p*=.009. The CHR group showed the highest percentage of motor delays with 16.5% of the CHR group reporting some or severe motor delays, and 100% of individuals reporting severe motor delays were also members of the CHR group. The frequency of engagement in motor hobbies (never, sometimes, often) were compared across the diagnostic groups (CHR, CSV, depression, anxiety); groups did not differ in terms of their engagement in fine motor hobbies, χ^2^(275)=8.94, *p*=.27.

### Convergent validity to motor performance

Composite scales were compared to performance on a finger tapping task performance. Within the CHR group, a repeated-measure general linear model compared motor speed across hands (dominant, non-dominant) by median and variance as the within-subject factor and the MAP-R subscales, respectively, as between-subject factors. Finger tapping performance related to the ***Motor Abnormalities*** subscale, *F*(71)=5.83, *p*=.016; higher self-reported ***Motor Abnormalities*** subscale related to lowered median taps on both the dominant (*r*_*partial*_= −.23) and non-dominant hands (*r*_*partial*_ = −.18). Finger tapping performance also related to the *Coordination Abnormalities* sub-facet, *F*(71)=5.95, *p*=.017, such that increased *Coordination Abnormalities* sub-facet scores were related to lowered median taps on both the dominant (*r*_*partial*_= −.26) and non-dominant hands (*r*_*partial*_= −.22), *Figure 4*. Finger tapping did not relate to any other subscales or items, *p’s*>.13.

### Predictive and convergent validity to clinical features

Composite subscales totals related to the psychosis risk score (SIPS-RC) in a correlation. ***Motor Abnormalities*** subscale related to SIPS-RC scores, *r*(312)=.15, *p*=.008. *Coordination Abnormalities* sub-facet related to SIPS-RC scores, *r*(310)=.16, *p*=.006. *Dyskinesia* sub-facet related to SIPS-RC scores, *r*(308)=0.118, *p*=.04. ***Physical Activity*** susbscale related to SIPS-RC scores, *r*(284)=-.150, *p*=.01. For each of the independent items a limited set of Likert-responses were compared across groups using a general linear model. SIPS-RC did not relate to infant delays (*p*=.23) or frequency of fine motor hobbies (*p*=.63).

## Discussion

The MAP-R scale showed a replicable structure that grouped items into ***Motor Abnormalities*** and ***Physical Activity*** subscales. ***Motor Abnormalities*** further divided into *Coordination Abnormalities* and *Dyskinesia* subscales. Each of these factors was of significant interest to understanding, identifying, predicting, and treating early psychosis, and so a motor scale that differentiates distinct motor categories has significant potential. Further, these subscales discriminated individuals at CHR from clinical groups (anxiety and depression) and a community sample of CSV. This discriminant validity suggested that this scale has sensitivity to psychosis-risk over issues related to general psychopathology, which would also appear in clinical samples in general. There was support for convergent validity; the general motor abnormality scale and the coordination scale were related to finger tapping performance. Additionally, these scales related to a validated measure of risk for psychosis indicating potential predictive validity of this scale to be relevant to psychosis course and risk for conversion to psychosis. Collectively this scale may provide unique insight into early risk for psychosis by examining motor abnormalities, which is largely under-utilized in current approaches to psychosis risk and may reflect a novel target for risk treatment.^31^

The exploratory and validation analytic dataset factor analyses suggested that items of the scale could be grouped into two subscales (***Motor Abnormalities*** and ***Physical Activity*** subscales) with 2 sub-facets (*Coordination Abnormalities, Dyskinesia*) in the exploratory analytic dataset.^22^ Scale structure was consistent across independent data points in the analytic datasets providing converging evidence grouping the same items into the same subscale structure, providing increased confidence in the stability of these subscale structures.^67^ Additionally, these scales showed similar reliability across the exploratory and validation analytic datasets further suggesting the stability of these subscales. It is also notable that both the exploratory and the validation analytic datasets reflected a diverse community sample, which should both increase our sensitivity to individual differences and increase the generalizability of the current scale structures.^70^ The current paper identified three scales that may reflect distinct vulnerabilities/mechanisms/circuits that are central to the etiology of psychosis, and reflected by motor abnormalities.^22^ For example, motor behaviors ranging from hippocampal vulnerability, BDNF, and neurogenesis in the case of sedation/physical activity,^27^ to cerebellar-thalamic dysfunction for poor coordination,^18,21^ balance and ataxia,^10,21,43,71^ to aberrant dopamine.^9,15,34,46,47^ Finally, the validation analyses provided initial evidence of the utility of these scales.

The validation analytic dataset was a diverse community sample^70^ that included CSV as well as individuals with psychiatric diagnoses including major depression disorder and anxiety disorders (which are associated with some degree of motor abnormalities, such as psychomotor slowing or psychomotor agitation).^72,70^ In these analyses, the CHR group showed significantly more motor abnormalities, coordination abnormalities, and dyskinesia indicating that these scales are particularly sensitive to the presence of psychosis risk, beyond general psychopathology or general psychomotor agitation/slowing, which may have been present in depression and anxiety. In contrast, the ***Physical Activity*** subscale did not discriminate between the CSV and the CHR group. However, the ***Physical Activity*** subscale was sensitive to the presence of general psychopathology as the psychiatric groups did not differ from each other, but differed from the CSV sample.^61,72^ It is notable that both the ***Motor Abnormalities*** subscale and the ***Physical Activity*** subscale distinguished other anxiety and depression from CSV. This sensitivity to other psychopathology suggests that this scale may also have some utility in examining motor abnormalities in depression and anxiety. In summary, the scales showed both a specific relationship to psychosis risk and a general sensitivity to discriminate individuals with psychopathology from a CSV group. However, this self-report measure relied on the patient’s insight and, as a result, may not reliably reflect actual motor behavior.^73–75^ To this end we have further validated these scales against a measured motor behavior.

The finger tapping task has been known to reflect underlying disturbances in the function of underlying neurocircuitry of motor systems.^30,76,77^ This task is particularly probative of cortico-cerebellar interactions that govern the execution of sub-second responses, but also motor slowing generally. Among the subscales examined, the coordination subscale was the most relevant scale related to cortico-cerebellar function,^76^ and was related to performance on the finger tapping task. Critically, this result suggests that individuals at CHR were able to faithfully report on their coordination in a way that reflected independent measures of motor behavior. Although the ***Motor Abnormalities*** subscale related to the performance on this task, this was driven by the coordination abnormalities items.

All of the MAP-R subscales related to the calculated risk for developing psychosis-a composite score that weights the predictive features of clinical high-risk for psychosis (i.e., SIPS-RC score).^64^ Calculated risk scale serves as an estimation of the likelihood of an individual to convert to psychosis, but it also leverages critical features of early clinical risk, e.g., symptom severity and decreased global function.^62,64^ Taken together, the relationship of these subscales for calculated risk may reflect the relevance of ***Motor Abnormalities*** subscale to risk for conversion to psychosis, as estimated by calculated risk for psychosis, consistent with previous research.^1,37^

Despite the promise of the MAP-R subscales to reflect theoretical factors, there were two items that did not map onto any particular subscale-motor developmental delays and engaging in fine motor skill hobby items. The motor developmental delays item distinguished between clinical groups with a larger proportion of individuals in the CHR group who endorsed minor motor developmental delays,^3,78^ and endorsements of major motor developmental delays were made up entirely of individuals at CHR. The motor delay item was included in future versions of the MAP-R scale, but the fine motor hobby item will not be included in the final scale as it did it reflect group membership, clinical risk features, or motor performance.

This study included many strengths, but there were limitations that should be explored in future studies. Although the current paper identified three scales that may reflect distinct vulnerabilities/mechanisms/circuits, we did not directly assess mechanistic ties and specificity in the current study. Further, we omitted additional motor behaviors such as gestures and catatonia that are likely to tap into other distinct vulnerabilities.^16,71,79–85^ Finally, in the absence of longitudinal data, it is unknown whether motor abnormalities relate to clinical course or ultimate conversion to psychosis.

The MAP-R scale fills a gap in current self-report and clinical interviews that assess clinical risk as these measures rely on a limited set of items on motor abnormalities, while the current scale includes diverse items that include abnormalities. This scale allows researchers to assess motor phenotypes without expensive equipment or expertise, and can be used in many different psychosis assessment settings as a low-cost assessment of mechanisms underlying emerging symptoms of psychosis. As a result the MAP-R Scale may be a beneficial tool in CHR screening in clinical practice and has the potential to be an important target for clinical trials.^31^

## Data Availability

Data is available upon request to PIs.

## Acknowledgments

This work was supported by the National Institutes of Mental Health (MH094650, MH112545, MH103231, MH094650, VM; 5R01MH112613-03, 3R01MH112613-02S1, and 5R01MH112613-02, LME; 5R01MH112612-03, 5R01MH112612-02, and 1R01MH112612-01). We have no conflicts to disclose.

---

**Supplementary Figure 1.**
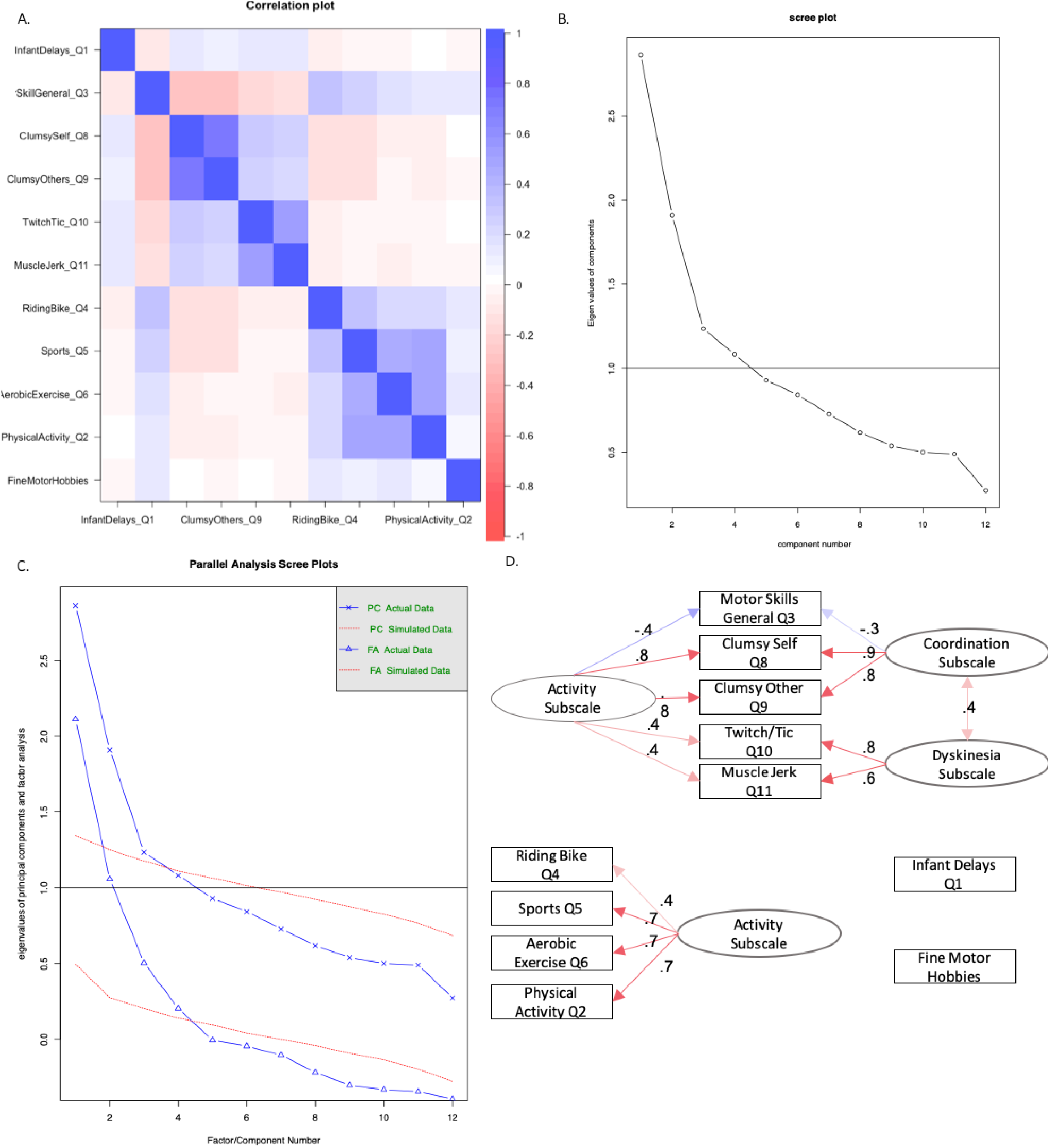
Factor analyses and data structure for the exploratory analytic dataset: A. Depicts the intercorrelation structure of the items, B. Depicts the Cattell scree plot of the intercorrelation matrix, C. Depicts the factor analyses compared to simulated data in a scree plot, D. Depicts the factor structure for both a 2 factor (***Motor Abnormalities*** and ***Physical Activity Scales***) and three factor solution (*Dyskinesia Abnormalities sub-facets, Coordination abnormalities sub-facets*, and ***Physical Activity Scale***)

